# LED transillumination of the chest and abdomen in the diagnosis of neonatal conditions

**DOI:** 10.1101/2021.08.29.21262761

**Authors:** José Abreu, Michelle Baldacci, Gladys Tarenzi, Karla Villegas

**Affiliations:** Universidad de Carabobo

**Keywords:** LED transillumination, neonate, pneumothorax, pneumoperitoneum, NICU

## Abstract

Air leak syndromes, such as pneumothorax and pneumoperitoneum, are commonly diagnosed in Neonatal Intensive Care Units. However, this finding is usually detected with radiographic studies when the neonate is in poor condition. LED transillumination is an alternative method that allows a fast, inexpensive, and non-invasive diagnosis. **We aimed** to evaluate the clinical utility of LED transillumination in neonates by comparing the results to chest and abdomen radiograph as reference test.

**Methodology:** diagnostic and prospective study that included 56 neonates. We used a simple and inexpensive handcrafted LED transilluminator to evaluate the chest and abdomen of patients, blind to the final diagnosis. The findings were compared to the radiographs.

**Results:** the study presented 3 true positive pneumothorax cases (sensibility 100%, 95%CI 29.24-100; specificity 100%, 95%CI 93.28-100), 1 true positive pneumoperitoneum case (sensibility 100%, 95%CI 2.50-100; specificity 100%, 95%CI 93.51-100), 10 true positive and 5 false negatives cases of gastric dilation (sensibility 66.67%, 95%CI 38.38-88.18; specificity 100%, 95%CI 91.19-100; negative likelihood ratio 0.33, 95%CI 0.16-0.68) and 4 true positive cases of dilation of bowel loops (sensibility 100%, 95%CI 39.76-100; specificity 100%, 95%CI 93.02-100).

**Discussion:** LED transillumination is a safe, inexpensive, and easy-to-use clinical tool that may be employed to promote early intervention during severe cases of air leak syndrome that require immediate action, especially in low-income and rural areas. Further research with larger sample sizes is required to properly define the efficacy and usefulness of LED transillumination in routine use.

## INTRODUCTION

Since the implementation of Neonatal Intensive Care Units (NICU), the care of critically ill newborns has advanced significantly by providing an environment where various therapeutic techniques are performed safely by specialized personnel to guarantee a better quality of life and life expectancy for the neonate. The use of mechanical ventilation (MV) has saved the lives of countless neonates in NICUs, a place where respiratory pathologies like acute respiratory distress syndrome (ARDS) represent a common problem.^1^ At the same time, MV has increased the incidence of pneumothorax (PT), a life-threatening complication characterized by the presence of air in the pleural space due to alveolar rupture.^2–4^ Pneumoperitoneum (PP), characterized by the presence of free air in the peritoneal cavity, is also gaining importance in neonates with necrotizing enterocolitis (NEC) or receiving MV.^5–7^

Among neonates, PT is associated with male sex, prematurity, respiratory diseases (ARDS, pulmonary hypoplasia, pulmonary interstitial emphysema, meconium aspiration syndrome), trauma, positive pressure ventilation,^3,8^ and reanimation manoeuvrers.^9^ It may develop spontaneously in 1-2% of newborns^3,7^ and up to 40% of neonates receiving MV as a result of volutrauma.^2,10^ The clinical presentation is initially asymptomatic; if the condition worsens, it is followed by signs of respiratory distress and gasometric alterations.^2,3,11^ PT diagnosis usually requires a chest radiograph showing radiolucency, pulmonary collapse, hemidiaphragm descent in the affected hemithorax, and contralateral mediastinal shift.^4^ If a radiographic study cannot be performed, minimal thoracostomy can be done with a small-calibre catheter connected to a water seal to demonstrate air leakage; however, this may result in iatrogenic PT, ineffective drainage, pain, and other complications.^12^ Upon confirmation, treatment consists of basic support measures and watchful waiting or placement of a chest tube for air drainage if the case warrants it.^3,13^

In newborns, PP is a complication of gastrointestinal perforation that often requires immediate surgical intervention.^5,6,14-17^ Prematurity and low birth weight are important risk factors due to their relation with NEC and intestinal immaturity.^14,18^ Abdominal radiographs in patients with NEC may reveal ileus, dilated bowel loops, pneumatosis intestinalis, pneumatosis portal, ascites, and PP.^14,16,19^ Treatment of NEC varies according to the stage of the disease, relying on bowel rest, intravenous alimentation, and broad-spectrum antibiotic therapy in the early stages as well as surgical intervention with drainage in case of peritoneal air leakage.^14^

The diagnosis of PT and PP usually occurs when the neonate’s condition is grave,^11^ favouring the development of severe complications such as intraventricular hemorrhage^9,20,21^ and peritonitis,^14,19^ respectively. For this reason, the application of practical methods aimed at rapid detection is required, especially in rural or low-income areas without easy access to imaging studies. Transillumination, which consists of the direct application of visible light through the skin to detect the presence of air or liquid,^22^ provides a non-invasive, fast, and cheap diagnosis for neonatal air leak syndromes.^17,23–26^

The traditional transilluminator used in neonates consists of a light source (usually halogen or incandescent) transmitted through a fibre optic cable.^17,24,25^ Such a device was employed by Buck et al. (1977) to diagnose thoracic, abdominal, cephalic, and cervical pathologies in neonates. It was utilized to identify PT, pneumomediastinum, pneumopericardium, and diaphragmatic herniation, detecting as low as 10cc of free air. They also reported gastric dilation (GD) and dilated bowel loops (DBL) in non-necessarily ill neonates.^24^ However, a case report by Murki et al. (2016) described the successful use of a LED (light-emitting diode) torch to detect PT in a preterm neonate receiving MV, opening the doors for LED transillumination.^27^

Considering the late diagnosis of PT^11^ and its potentially life-threatening complications,^9,20^ some authors consider the use of thoracic transillumination for a faster diagnosis.^21,24,26,28^ Also, the need for urgent surgical intervention in patients with PP,^5,6,14–17^ the frequent exposure to radiation in NICUs,^5,21^ the need for radiological protection,^29^ and the lack of new research about neonatal transillumination motivated our team to study the thoracic and abdominal findings of LED transillumination.

## METHODOLOGY

The study design was diagnostic and prospective. We evaluated the diagnostic utility of thoracic and abdominal transillumination with an LED device. We achieved this by correlating transillumination (index test) and radiographic findings (reference standard) in newborns from the NICU from Central Hospital of Maracay between 26 April and 23 August 2019.

We obtained written informed consent from the parents or caretakers of the neonates before enrolment. The study included neonates that fulfilled the following criteria: ≤28 days of age, available clinical history with the clinical-epidemiological data required for the study, and cases tested with transillumination and radiographs of the thorax and abdomen within 24 hours.

The transillumination device employed during the study was built by the researchers Jose Abreu and Michelle Baldacci, and it consists of a red LED bulb connected to rechargeable batteries. Most components were recycled (cables, case, and batteries). Compared with a traditional fiberoptic transillumination device, our LED transilluminator is very simple and inexpensive to make.

The thoracic transillumination data were recollected via direct observation, presenting three diagnostic possibilities: “positive in right hemithorax”; “positive in left hemithorax”; and “negative.” The findings for abdominal transillumination were obtained via direct observation, with five diagnostic possibilities: “positive in right upper quadrant”; “positive in left upper quadrant”; “positive in right lower quadrant”; “positive in left lower quadrant”; and “negative.” Data from thoracic and abdominal radiographs were obtained via direct observation from the NICU’s imagenology database. The researchers were blind to radiographic diagnosis before and during transillumination.

Before applying the index test, we tried to darken the surroundings as much as possible. We covered the LED bulb with a disposable transparent plastic sheet or cellophane to avoid cross-contamination between neonates;^22^ then, the bulb was applied above and below the nipple,^24,28^ in the axillary line, and under the subcostal region.^28^ This was repeated and compared with the contralateral side. We considered a hemithorax transillumination of ≥3 centimetres in diameter around the bulb as positive transillumination suggestive of PT. We also investigated the presence of other thoracic air leak syndromes. Pneumomediastinum is characterised by the visualization of cardiac pulsations through transillumination. The diagnosis of pneumopericardium is characterised by a bright transillumination around the pericardial sac in the third or fourth intercostal space with the light beam pointed towards the xiphoid appendix; cardiac pulsations are not necessarily evident.^22^

We applied the LED transilluminator in both flanks, the periumbilical region, and the epigastrium during the abdominal evaluation. Complete abdominal transillumination was interpreted as PP. Complete abdominal transillumination with visualization of the liver and spleen was diagnosed as ascites. Well-defined transillumination in the left upper quadrant suggests GD, and focal transillumination in any quadrant indicates DBL.^24^

The database was created in Microsoft Excel and processed with Epi Info™ version 7.2.3.1 for statistical analysis. During the statistical analysis, we used absolute and relative frequency data, 95% confidence interval, mean, standard deviation, median, range, and we performed a statistical comparison between groups with the Chi-square test to identify significant differences with the p-value obtained through the ANOVA test, with a value of p ≤0.05 considered as statistically significant. The reporting of our study followed the STARD (Standards for Reporting of Diagnostic Accuracy Studies) 2015 guidelines.^30^

The research complied with international (Helsinki Declaration 2013), national (Código de ética para la vida [Code of Ethics for Life]; article 41 of the LOPPNNA), and institutional (Bioethics Committee of the Central Hospital of Maracay) ethical criteria. We preserved the confidentiality of the neonates and their representatives, did not include patients without the written informed consent of their caretaker and did not apply unjustified or harmful procedures to the physical and mental integrity of the newborns.

## RESULTS

We included 56 neonates (28 males and 28 females) from the NICU of the Central Hospital of Maracay between 26 April and 23 August 2019 in the study. Initially, we considered eligibility criteria among a total of 90 NICU patients, of whom 34 did not meet the inclusion criteria. As shown in Figure 1, 2 patients exceeded the 28 days of age, 13 did not grant informed consent, 12 did not have a radiographic study within 24 hours after the examination, and 7 did not have their clinical history available at the time of the evaluation. The remaining 56 cases met the inclusion criteria.

**Figure 1.**
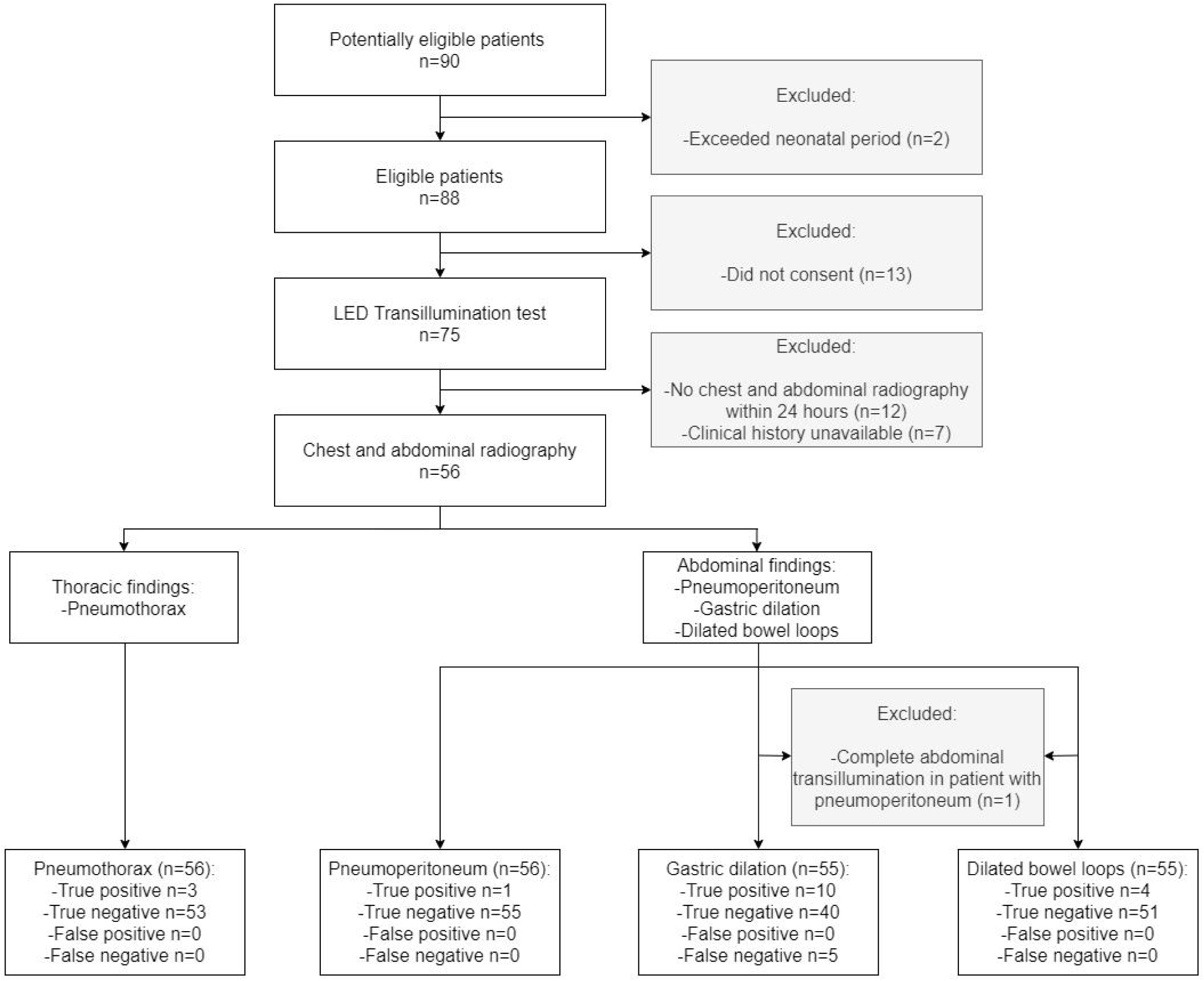
Flow diagram. The study compared transillumination with a LED device (index test) with chest and abdominal radiographs (reference test) for pneumothorax, pneumoperitoneum, gastric dilation, and dilated bowel loops.

**Figure 2.**
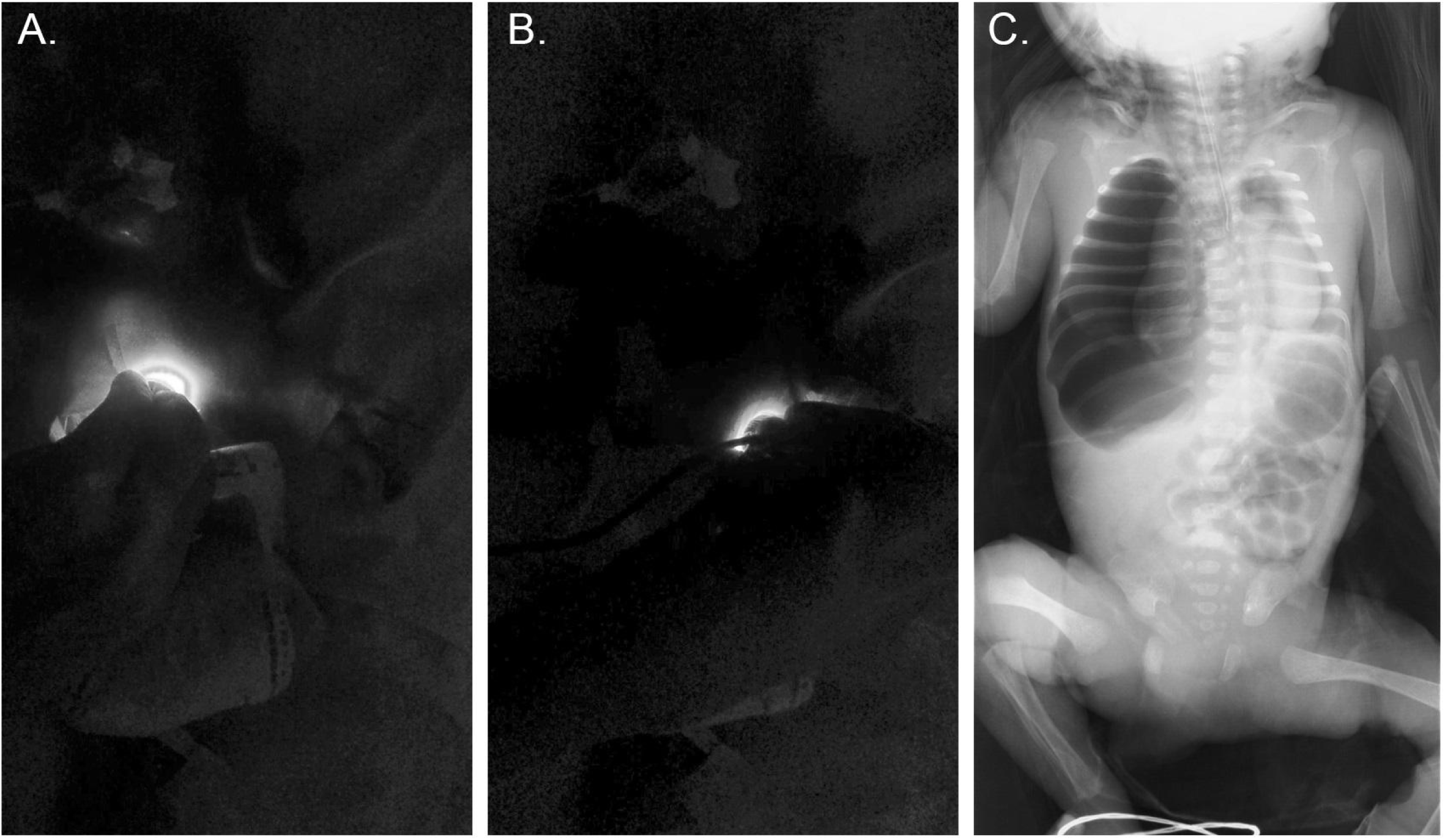
Positive transillumination in the right hemithorax, compatible with pneumothorax (A). Negative transillumination in the contralateral hemithorax (B). The diagnosis was soon confirmed with a chest radiograph (C).

During the study, the mean gestational age was 32.76 (±2.93) weeks. Most patients were moderate preterm (21 [37.5%], 95%CI 24.92-51.45) and very preterm infants (19 [33.93%], 95%CI 21.81-47.81), as shown in Table 1. Mean birth weight was 1822.59 (±465.99) grams and 47 neonates had adequate weight for gestational age (83.93%, 95%CI 71.67-92.38). Neonates presented a mean one-minute APGAR score of 6.41 (±1.25) and a five-minute score of 7.68 (±0.72).

**Table 1.** Mean, standard deviation, frequency, and 95% confidence interval of epidemiological and perinatal characteristics of neonates.

|  |  | Cases (n=56) |  |
| --- | --- | --- | --- |
| | | n(%) or $\bar{x}$ | 95%CI or ±SD |
| **Sex** |  |  |  |
|  | Male | 28(50) | 36,34-63,66 |
|  | Female | 28(50) | 36,34-63,66 |
| **Gestational age (weeks)** |  | 32.76 | 2.93 |
| **Gestational age classification** |  |  |  |
|  | Extremely preterm | 3(5,36) | 1,12-14,87 |
|  | Very preterm | 19(33,93) | 21,81-47,81 |
|  | Moderate preterm | 21(37,5) | 24,92-51,45 |
|  | Late preterm | 7(12,5) | 5,18-24,07 |
|  | Full term | 6(10,71) | 4,03-21,88 |
| **Birth weight (grams)** |  | 1822.59 | 465.99 |
| **Weight classification** |  |  |  |
|  | SGA | 5(8,9) | 2,96-19,62 |
|  | AGA | 47(83,93) | 71,67-92,38 |
|  | LGA | 4(7,14) | 1,98-17,29 |
| **One-minute APGAR** |  | 6.41 | 1.25 |
| **Five-minute APGAR** |  | 7.68 | 0.72 |
| **Maternal age (years)** |  | 24.93 | 7.57 |
| **Prenatal visits** |  | 3.89 | 2.82 |

95%CI: 95% confidence interval; SD: standard deviation; Extremely preterm: <28 weeks; Very preterm: $\geq 28$ - $\leq 32$ weeks; Moderate preterm: $\geq 32$ - $\leq 35$ weeks; Late preterm: $\geq 35$ - $\leq 37$ weeks; Full term: $\geq 37$ weeks; SGA: small for gestational age; AGA: adequate for gestational age; LGA: large for gestational age.

As shown in Table 2, ARDS was the most frequent associated condition with 33 cases (58.93%, 95%CI 44.98-71.90). Stage IA NEC was diagnosed in 12 neonates (21.43%, 95%CI 11.59-34.44). There were 22 cases of sepsis (39.29%, 95%CI 26.50-53.25), represented by 18 cases of early neonatal sepsis (32.14%, 95%CI 20.29-45.96).

**Table 2.** Frequency and 95% confidence interval of the clinical findings.

|  | Cases (n=56) |  |
| --- | --- | --- |
|  | n(%) | 95%CI |
| **Respiratory pathologies** |  |  |
| ARDS | 33(58,93) | 44,98-71,90 |
| TTN | 5(8,93) | 2,96-19,62 |
| MAS | 2(3,57) | 0,44-12,31 |
| Pneumonia | 10(17,86) | 8,91-30,40 |
| RD-BH | 4(7,14) | 1,98-17,29 |
| None | 2(3,57) | 0,44-12,31 |
| **Gastrointestinal pathologies** |  |  |
| IA NEC | 12(21,43) | 11,59-34,44 |
| IB NEC | 1(1,79) | 0,05-9,55 |
| Gastroschisis | 1(1,79) | 0,05-9,55 |
| Jejunoileal atresia | 1(1,79) | 0,05-9,55 |
| None | 41(73,21) | 59,70-84,17 |
| **Sepsis** |  |  |
| Early-onset sepsis | 18(32,14) | 20,29-45,96 |
| Late-onset sepsis | 3(5,36) | 1,12-14,87 |
| Septic shock | 1(1,79) | 1,12-14,87 |
| None | 34(60,71) | 46,75-73,50 |
| **Ventilation time** |  |  |
| MV | 7(12,50) | 5,18-24,07 |
| nCPAP | 39(69,64) | 55,90-81,22 |
| None | 10(17,86) | 8,91-30,40 |
| **Diet** |  |  |
| Breast milk | 16(28,57) | 17,30-42,21 |
| Nothing by mouth | 40(71,43) | 57,79-82,70 |

95%CI: 95% confidence interval; ARDS:acute respiratory distress syndrome; TTN: transient tachypnea of the newborn; MAS: meconium aspiration syndrome; RD-BH: respiratory distress secondary to brain hypoxia; NEC: necrotizing enterocolitis; MV: mechanical ventilation; nCPAP: nasal continuous positive airway pressure.

There were only ten neonates without ventilatory support (17.86%, 95%CI 8.91-30.40). The use of nCPAP was frequent (39 [69.64%], 95%CI 55.90-81.22), and the mean ventilation time was 2.41 (±2.26) days. There were 40 neonates receiving nothing by mouth (71.43%, 95%CI 57.79-82.70), and the remaining 16 received breast milk (28.57%, 95%CI 17.30-42.21).

The mean time between transillumination and radiograph was 5.18 (±6.66) hours, with a median of 3 hours (range, 0-23 hours).

There were three positive hemithorax transillumination findings compatible with PT, as evidenced in Figure 2. Two cases were considered positive due to evident irregular illumination of the affected hemithorax and the remaining due to increased transillumination around the bulb in contrast with the contralateral hemithorax. The diagnosis of PT was confirmed with a chest radiograph, obtaining 3 true positives and 53 true negatives (Table 3).

**Table 3.**
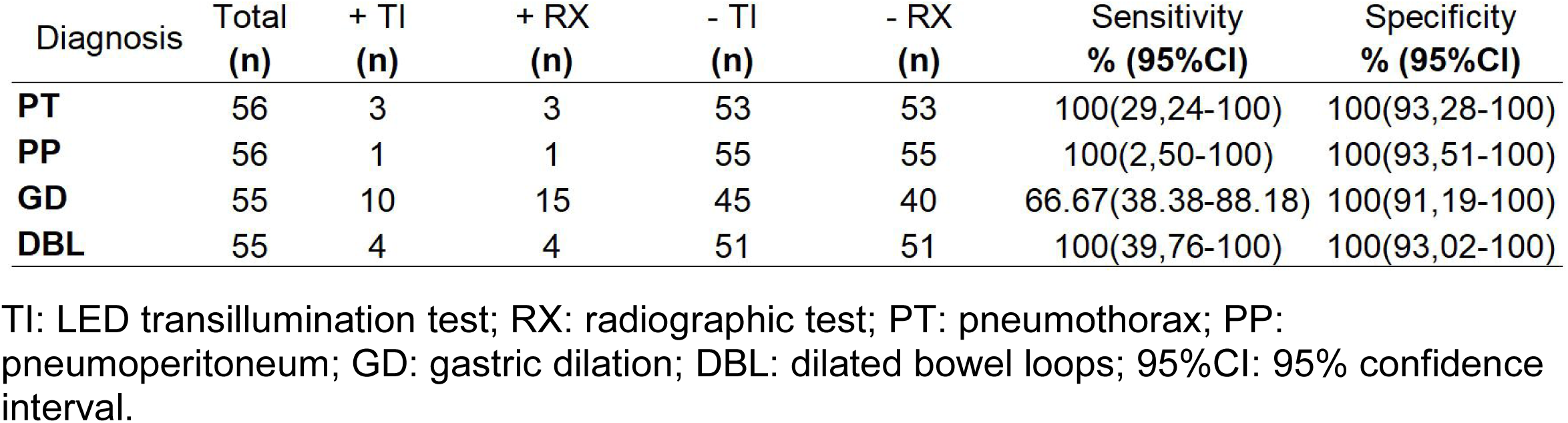
Sensitivity, specificity, and 95% confidence interval of LED transillumination when compared to the radiographic diagnosis of pneumothorax, pneumoperitoneum, gastric dilation, and dilated bowel loops.

| Diagnosis | Total<br>(n) | + TI<br>(n) | + RX<br>(n) | - TI<br>(n) | - RX<br>(n) | Sensitivity<br>% (95%CI) | Specificity<br>% (95%CI) |
| --- | --- | --- | --- | --- | --- | --- | --- |
| **PT** | 56 | 3 | 3 | 53 | 53 | 100(29,24-100) | 100(93,28-100) |
| **PP** | 56 | 1 | 1 | 55 | 55 | 100(2,50-100) | 100(93,51-100) |
| **GD** | 55 | 10 | 15 | 45 | 40 | 66.67(38.38-88.18) | 100(91,19-100) |
| **DBL** | 55 | 4 | 4 | 51 | 51 | 100(39,76-100) | 100(93,02-100) |

TI: LED transillumination test; RX: radiographic test; PT: pneumothorax; PP: pneumoperitoneum; GD: gastric dilation; DBL: dilated bowel loops; 95%CI: 95% confidence interval.

One neonate presented complete abdominal transillumination. It was compatible with PP because it was bright, and neither the hepatic silhouette nor spleen could be distinguished. The abdominal radiograph confirmed the diagnosis (Figure 3). Considering that the complete abdominal transillumination could have obstructed the visualization of GD or DBL, which did not occur in this particular case, we decided not to include this patient during the calculation and analysis of other abdominal findings.

**Figure 3.**
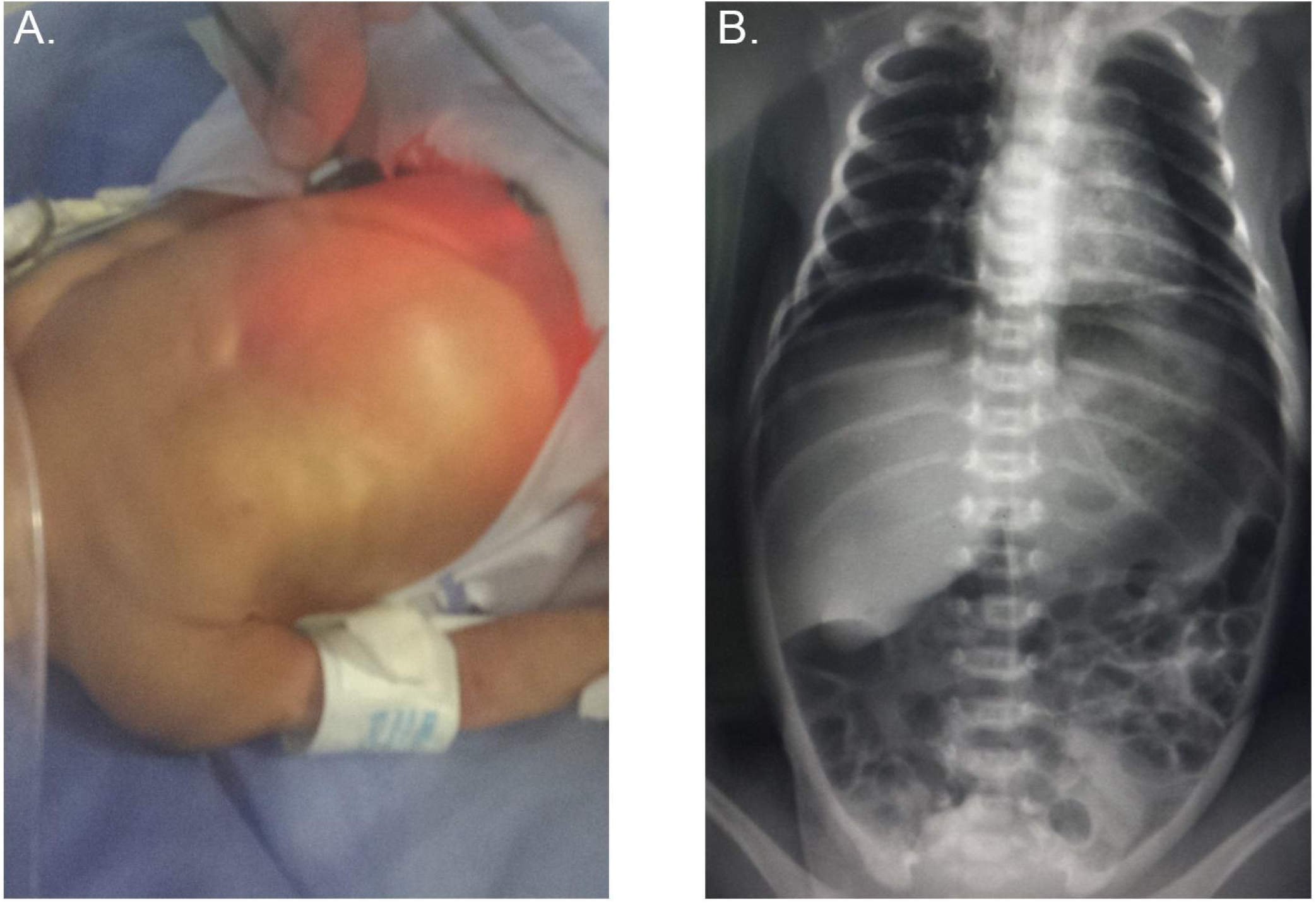
Complete abdominal transillumination suggestive of pneumoperitoneum (A). The corresponding radiograph, confirming the diagnosis (B).

There were ten neonates with true positive GD identified by LED transillumination and five false negatives (sensitivity 66.67%, 95%CI 38.38-88.18; specificity 100%, 95%CI 91.19-100; negative likelihood ratio 0.33, 95%CI 0.16-0. 68). GD findings were characterized by an intense, well-defined transillumination located in the upper left quadrant (see Figure 4).

**Figure 4.**
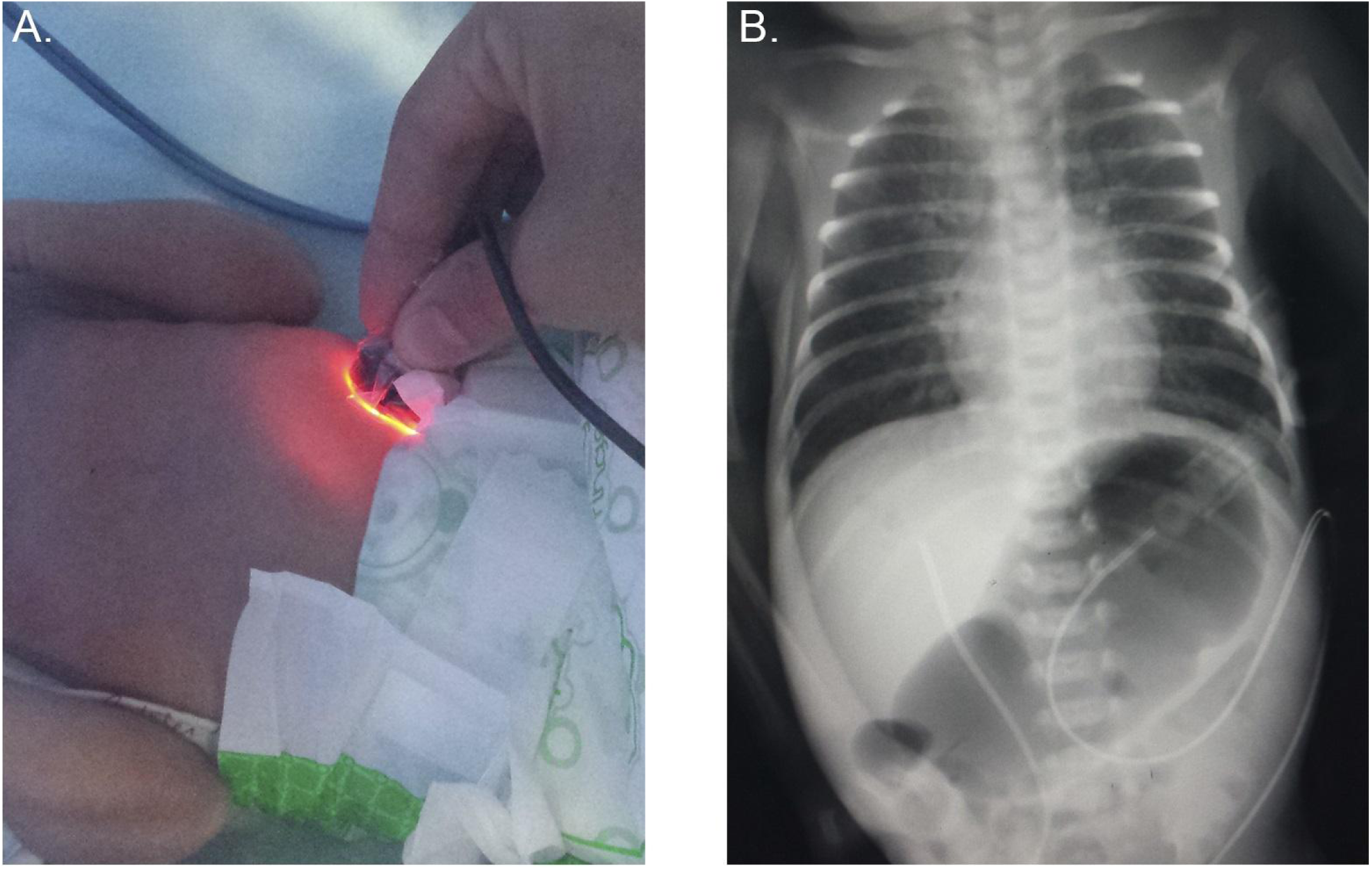
Bright and well-defined in the upper left quadrant, congruent with gastric dilation (A). The radiograph showed evident dilation, confirming the diagnosis (B).

Findings compatible with DBL (sensitivity of 100%, 95%CI 39.76-100; specificity of 100%, 95%CI 93.02-100) were characterized by being located in any abdominal quadrant. DBL presents a dim and diffuse transillumination (see Figure 5).

**Figure 5.**
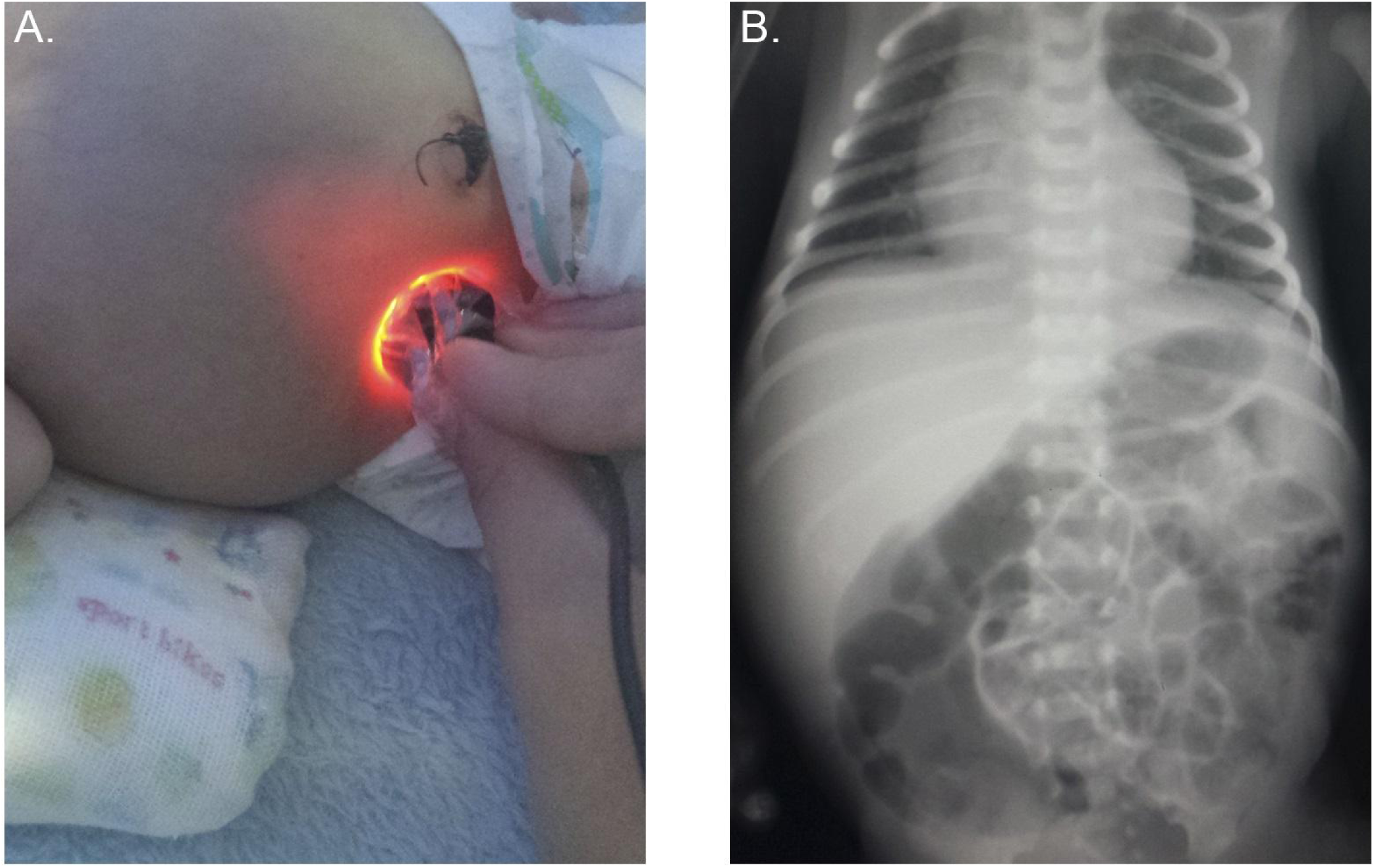
Dim transillumination in the right lower quadrant, suggestive of dilated bowel loops (A). The abdominal radiograph confirmed this finding, showing bowel dilation in the same area (B).

Two cases stand out among the neonates that did not meet all the inclusion criteria: a case of positive transillumination in the right hemiabdomen suggestive of ascites due to good visualization of the hepatic border, which could not be confirmed due to the death of the neonate. The other case was a neonate with duodenal atresia that exceeded the neonatal period and presented positive transillumination in the upper hemiabdomen.

No adverse events related to LED transillumination occurred during the study, such as skin burns or other injuries.

## DISCUSSION

Our main discovery is that LED transillumination can diagnose the presence of multiple thoracic and abdominal findings in a blind comparison with our reference test (chest and abdominal radiograph). To our knowledge, this is the first diagnostic study that has explored LED transillumination for this application. In contrast with traditional devices, the use of LEDs in transilluminators minimizes the risk of burns and may significantly lower the cost of the device. Developing and crafting the transilluminator used during the study did not require advanced technical knowledge. Except for the LED bulb, all the materials were recycled from old electronic devices.

The main cause of neonatal morbidity were respiratory pathologies, observed in 54 neonates (96.43%, 95%CI 87.69-99.56), followed by 22 cases of sepsis (39.29%, 95%CI 26.5-53.25), and 15 neonates with gastrointestinal pathologies (26.79%, 95%CI 15.83-40.3). These findings are similar to those reported in other healthcare centres in our country.^1^ Ventilatory support was frequent during our study (46, 82.14%), which was expected in a NICU with such a high incidence of respiratory pathologies.

Thoracic transillumination with a LED device allowed the diagnosis of 3 true positive cases of PT and 53 true negative cases. There were no false positives in our cohort; however, Kuhns et al.^23^ reported a 4% false positive rate. The sensitivity (100%, 95%CI 29.24-100) is compatible with the results reported by Buck et al.^24^ (96.49%, 95%CI 87.89-99.57) and Wyman et al.^26^ (94%, 95%CI 83.45-98.75), who did not report true negative or false positive cases. Our study included every neonate that fulfilled the inclusion criteria, in contrast with previous studies that specifically targeted neonates that were suspicious of air leak syndromes.

Evidencing a positive transillumination in patients with PT required placing the transilluminator in the subcostal region (as shown in Figure 2A). The reason for this might be the LED bulb we employed, which was circular and too large to fit between the ribs of neonates, preventing the adequate transmission of light.

Similar to a diagnostic study by Buck et al.,^24^ abdominal transillumination was helpful during the diagnosis of GD, DBL, and PP; however, there were no confirmed cases of ascites. Unlike the cases of PP reported by Togari et al.,^17^ Demaret et al.,^31^ and El-Matary et al.,^32^ the neonate in our study did not require surgical intervention and resolved spontaneously like a case reported by Gupta et al.^33^

There were several limiting factors during our research. The illumination of the area was a major constraint, as it was not possible to adjust its intensity most of the time. For this reason, it was not possible to apply the index test with the same degree of illumination to all the neonates in the area; some incubators had a blue curtain which allowed to reduce the luminosity inside the incubator. The imagenology department did not follow a fixed schedule in the NICU, so radiographs were taken at different times of the day. This may have impacted the effectiveness of the index test in certain cases. For example, when discarding radiographs taken three hours or more after transillumination, the false negative GD cases are reduced from five to one. Occasionally, they were not taken for more than 24 hours; this was especially troubling between April and July 2019, when our country had severe problems with the electric power system. A few radiographs were lost during this period, which reduced our sample size.

The study had a reduced application period (April-August), resulting in a small sample. PT and PP had a low incidence during our research, and they were difficult to detect because they usually require immediate resolution. The use of adhesives, electrodes, and other materials may have hindered the correct visualization of transillumination, but this did not lead to false diagnoses.

We recommend performing a study with larger sample size. Transillumination should be performed prior to radiography within a three-hour window. The use of adjustable light and transparent adhesives in neonatal areas is recommended; nonetheless, it is not required for the application of LED transillumination.

In conclusion, LED transillumination is a safe, inexpensive, and easy-to-use clinical tool that may be employed to promote early intervention during severe cases of air leak syndrome that require immediate action, especially in low-income and rural areas. Further research with larger sample sizes is needed to properly define the efficacy and usefulness of LED transillumination in routine use.

## Data Availability

The data collected during the study is stored in the personal computer of the authors, as well as in a private cloud-based storage system. If required, such data is readily-available for revision.

